# Patient characteristics, clinical care, resource use, and outcomes associated with hospitalization for COVID-19 in the Toronto area

**DOI:** 10.1101/2020.12.15.20248199

**Authors:** Amol A. Verma, Tejasvi Hora, Hae Young Jung, Michael Fralick, Sarah L. Malecki, Lauren Lapointe-Shaw, Adina Weinerman, Terence Tang, Janice L. Kwan, Jessica J. Liu, Shail Rawal, Timothy C.Y. Chan, Angela M. Cheung, Laura C. Rosella, Marzyeh Ghassemi, Margaret Herridge, Muhammad Mamdani, Fahad Razak

## Abstract

**Background:** Patient characteristics, clinical care, resource use, and outcomes associated with hospitalization for coronavirus disease (COVID-19) in Canada are not well described.

**Methods:** We described all adult discharges from inpatient medical services and medical-surgical intensive care units (ICU) between November 1, 2019 and June 30, 2020 at 7 hospitals in Toronto and Mississauga, Ontario. We compared patients hospitalized with COVID-19, influenza and all other conditions using multivariable regression models controlling for patient age, sex, comorbidity, and residence in long-term-care.

**Results:** There were 43,462 discharges in the study period, including 1,027 (3.0%) with COVID-19 and 783 (2.3%) with influenza. Patients with COVID-19 had similar age to patients with influenza and other conditions (median age 65 years vs. 68 years and 68 years, respectively, SD<0.1). Patients with COVID-19 were more likely to be male (59.1%) and 11.7% were long-term care residents. Patients younger than 50 years accounted for 21.2% of all admissions for COVID-19 and 24.0% of ICU admissions. Compared to influenza, patients with COVID-19 had significantly greater mortality (unadjusted 19.9% vs 6.1%, aRR: 3.47, 95%CI: 2.57, 4.67), ICU use (unadjusted 26.4% vs 18.0%, aRR 1.52, 95%CI: 1.27, 1.83) and hospital length-of-stay (unadjusted median 8.7 days vs 4.8 days, aRR: 1.40, 95%CI: 1.20, 1.64), and not significantly different 30-day readmission (unadjusted 8.6% vs 8.2%, aRR: 1.01, 95%CI: 0.72, 1.42).

**Interpretation:** Adults hospitalized with COVID-19 during the first wave of the pandemic used substantial hospital resources and suffered high mortality. COVID-19 was associated with significantly greater mortality, ICU use, and hospital length-of-stay than influenza.

## Introduction

The global coronavirus disease 2019 (COVID-19) pandemic has disrupted healthcare systems around the world. Studies in the United States, United Kingdom, and China report that patients hospitalized with COVID-19 have high rates of critical illness and mortality.^1–5^ Two small Canadian case series have described care for critically ill patients with COVID-19 and found that mortality was up to 25%.^6,7^ However, hospitalizations for COVID-19 in Canada are not well described, particularly outside of ICU. More than 80% of patients hospitalized for COVID-19 may not require intensive care.^1,4^ The features of COVID-19 hospitalizations in Canada may differ from other countries due to differences in populations, public health, and health care systems. Case fatality rates for COVID-19 vary dramatically worldwide.^8^ In Canada, COVID-19 has disproportionately affected urban and suburban ethnic minority populations,^9^ and residents of Long-Term Care facilities^10,11^.

Seasonal influenza is a useful comparator to assess the severity of illness associated with COVID-19^12^ as it is another respiratory virus with high attendant morbidity and mortality, and it is familiar to the general public. The relative severity of COVID-19 and seasonal influenza remains controversial.^13^ The preponderance of evidence suggests that COVID-19 has greater morbidity and mortality but the magnitude of this difference remains uncertain.^13–18^ The purpose of this study was to describe patient characteristics, resource use, clinical care, and outcomes for patients hospitalized with COVID-19 at 7 hospitals in Toronto and Mississauga, Ontario, Canada. We compared hospitalizations for COVID-19 with hospitalizations for influenza and all other medical ward and intensive care unit (ICU) admissions.

## Methods

### Design and Setting

This retrospective cohort study was conducted at 7 large hospitals (5 academic and 2 community-based teaching hospitals) in Toronto and Mississauga, Ontario, participating in GEMINI, a hospital research collaborative.^19^ Research ethics board approval was obtained from all participating organizations.

### Data Collection

Administrative and clinical data were collected from hospital information systems for GEMINI as has previously been described in detail ^19,20^. Manual validation of a subset of GEMINI data found 98-100% accuracy compared to detailed medical record review.^20^ Patient demographics, hospital resource use, and outcomes were collected from hospitals as reported to the Canadian Institute for Health Information (CIHI) Discharge Abstract Database and National Ambulatory Care Reporting System. Additional clinical data, including laboratory test results, radiology tests, vital signs and in-hospital medication orders were extracted from hospital information systems. At one hospital, vital signs were manually abstracted only for patients with COVID-19 because they were not available through electronic extraction.

### Study population

We included all adult patients (greater than 18 years) discharged from an inpatient medical service or medical-surgical intensive care unit (ICU), including coronary care units, between November 1, 2019 and June 30, 2020. Medical services included all medical specialties (e.g. general medicine, cardiology, respirology, etc.). This captures all hospitalizations for COVID-19 and influenza and their medical complications but misses a small number of patients hospitalized for non-medical reasons (e.g. surgical, psychiatric, obstetrical) who were incidentally found to have COVID-19 or influenza and were not subsequently transferred to a medical service.

### Exposures

We categorized patients into three mutually-exclusive categories: COVID-19, influenza, and other medical admissions. We identified patients with COVID-19 based on International Statistical Classification of Diseases and Related Health Problems 10^th^ revision (ICD-10-CA) codes U07.1 (“COVID-19 diagnosis confirmed by a laboratory test”)^21^ and U07.2 (“COVID-19 diagnosed clinically or epidemiologically but lab results inconclusive, unavailable, or not performed”).^21^ We identified patients with influenza based on ICD-10-CA codes J09-J11, which an Ontario hospital study found to be 91% sensitive and 97% specific.^22^ Cases with both influenza and COVID-19 were categorized in the COVID-19 group. All admissions other than COVID-19 and influenza were considered for “other” conditions.

### Outcomes and Process Measures

We report in-hospital mortality and readmission to any medical or medical-surgical ICU service at any participating hospital within 7- and 30-days of discharge. To report readmission, we excluded patients who died and those discharged within the last 7-days and 30-days of the study period, respectively. We also report total hospital length-of-stay, emergency department (ED) length-of-stay, admission to the ICU, and ICU length-of-stay. We also describe use of thoracic computed tomography (CT) and in-hospital use of respiratory-acting antibiotics^23–25^ (see Appendix for details), anticoagulants, and systemic corticosteroids, as captured by medication orders after admission. These medications were selected because their prescribing may be associated with COVID-19,^26,27^ although the study period was prior to the publication of the RECOVERY dexamethasone trial results.^26^ We describe the use of invasive mechanical ventilation, renal replacement therapy (including both newly initiated and chronic hemodialysis and peritoneal dialysis), gastrointestinal endoscopy, and bronchoscopy identified using Canadian Classification of Health Interventions codes.

### Patient Characteristics

We report patient age, sex, residence in long-term care facility, and transfer from acute care hospital as reported by hospitals to CIHI. We categorized comorbid conditions based on ICD-10-CA codes using the Clinical Classification Software Refined (CCSR),^28,29^ the Charlson Comorbidity Index,^30^ and the Hospital Frailty Risk Score^31,32^. We report baseline laboratory test results and vital signs, defined as the first measurement that occurred between ED triage and 48 hours after admission.

### Statistical Analysis

We compared baseline characteristics between groups using standardized differences (SD), with SD>0.1 suggesting imbalance between groups.^33^ We compared unadjusted differences in clinical care, resource use and clinical outcomes between groups using chi-square tests for categorical variables, one-way ANOVA for continuous variables with a normal distribution, and Kruskall-Wallis tests for continuous variables with a non-normal distribution. We used multivariable regression to compare outcomes between groups after adjusting for patient age, sex, Charlson score, residence in long-term care, and admitting hospital. Poisson regression^34^ was used for mortality, readmission, and ICU admission and negative binomial regression was used for hospital length-of-stay and ICU length-of stay. Separate models were fit to compare COVID-19 to influenza and COVID-19 to other conditions.

### Subgroup and Sensitivity Analyses

First, we examined outcomes stratified by age groups. Second, we report patient characteristics and outcomes among the patients admitted to ICU. Third, we replicated all analyses after excluding COVID-19 diagnoses based on code U07.2 (patients with no laboratory confirmation). Finally, because participating hospitals are tertiary and quaternary care centres with large critical care units, we replicated analyses after including only patients admitted through the ED. This excluded interfacility transfers, which primarily involve patients transferred for critical care and might lead to an overestimation of illness severity.

## Results

During the study period there were 43,462 discharges, representing 34,782 unique patients. There were 1,027 discharges (3.0%) with COVID-19 (944 laboratory-confirmed), representing 972 unique patients. These represented 23.5% of all Ontario hospitalizations for COVID-19 (N= 4,373)^35^ during the study period. There were 783 discharges (2.3%) with influenza, representing 763 unique patients. Fewer than 6 patients had coexisting COVID-19 and influenza (exact value suppressed to limit risk of patient reidentification).

### Patient Characteristics

Patients with COVID-19 had a median age of 65 years (IQR 53, 79) which was similar to influenza (median 68 years, IQR 55, 80) and other conditions (median 68 years, IQR 55, 80, SD<0.1) (Table 1). Patients with COVID-19, compared to patients with influenza or other conditions, were more likely to be male (59.1% vs 50.8% and 56.1%, respectively, SD=0.17), more likely to have a Charlson score of zero (54.1% vs. 38.8% and 43.6%, respectively, SD=0.31), more likely to have intermediate or high hospital frailty risk score (37.2% vs 28.2% and 27.3%, respectively, SD=0.22), and more likely to reside in a long-term care facility (11.7% vs 4.5% and 2.9%, respectively, SD=0.34).

Hypertension (34.7%) and diabetes mellitus (27.7%) were common in patients with COVID-19 and occurred at similar rates in influenza and other conditions (Table 1). Neurocognitive disorders (including delirium and dementia) were more common in patients with COVID-19 compared to influenza and other conditions (16.9% vs. 13.4% and 10.1%, respectively, SD=0.20). Chronic obstructive pulmonary disease was more common in patients with influenza (12.3%) than with COVID-19 (5.4%) or other conditions (5.7%, SD=0.25).

Differences in baseline vital signs and laboratory test results are presented in Appendix Table 1. Presenting vital signs did not appear to differ meaningfully across groups, although patients with COVID-19 were more likely to require oxygen within 48 hours of hospitalization (46.0% vs. 40.1% and 21.3.6%, respectively, p <0.001). Patients with COVID-19 were more likely to have arterial blood gases (20.3% vs. 14.3% and 9.6%, p <0.001) and D-dimer measured (38.4% vs. 4.3% and 3.7%, respectively, p<0.001). Patients with COVID-19 had lower arterial pO2 (74.0 vs 81.5 and 108.0, respectively, p<0.001) but no significant difference in D-dimer values (p=0.22).

### Mortality and Readmission

Compared to patients with influenza and other conditions, patients with COVID-19 had significantly greater unadjusted in-hospital mortality (19.9% vs 6.1% and 5.9%, respectively, p<0.001, Table 2). Readmission within 7- and 30-days occurred in 4.4% and 8.6% of patients with COVID-19, respectively, which was not significantly different from influenza and other conditions.

Regression model results are reported in Table 3. After adjustment, patients with COVID-19 were more likely to die in hospital compared to patients with influenza (adjusted Relative Risk 3.47, 95% CI: 2.57, 4.67) and other conditions (aRR 3.58, 95% CI: 3.16, 4.06). There was no significant difference in 30-day readmission.

### Hospital Resource Use and Clinical Care

Compared to patients with influenza and other conditions, patients with COVID-19 had greater unadjusted ICU use (26.4% vs 18.0% and 23.5%, respectively, p<0.001) and hospital length-of-stay (median 8.7 days vs.4.8 days and 4.4 days, respectively, p<0.001) and shorter ED length-of-stay (median 8.7 hours vs. 21.1 hours and 13.6 hours, respectively p<0.001) (Table 2). COVID-19 patients were more likely to receive invasive mechanical ventilation (18.5% vs 9.3% and 9.7%, respectively) and renal replacement therapy (7.7% vs 5.5% and 4.3%, respectively, p <0.001) but were less likely to receive bronchoscopy or endoscopy (Table 2).

Patients with COVID-19 received at least one thoracic CT in 19.7% of admissions and at least one respiratory-acting antibiotic order in 71.6% of cases, which was similar to influenza and greater than other conditions (Table 2). Patients with COVID-19 received systemic corticosteroids in 16.7% of cases and received either warfarin or a direct-acting oral anticoagulant in 15.4% of cases.

After multivariable adjustment, patients with COVID-19 were more likely to use ICU than patients with influenza (aRR 1.52, 95% CI: 1.27, 1.83) and other conditions (aRR 1.21, 95% CI: 1.09, 1.34). Adjusted total hospital length-of-stay was greater for COVID-19 than influenza (adjusted rate ratio: 1.40, 95%CI: 1.20, 1.64, corresponding to an additional 1.45 days, 95%CI: 0.5, 3.2 days) and other conditions (aRR: 1.79, 95%CI: 1.66, 1.92, corresponding to an additional 4.12 days, 95% CI: 3.03, 5.52 days).

### Age-Stratified Outcomes

Age-stratified outcomes are presented in Table 4. Among patients with COVID-19 who were less than 50 years, 50-75 years, and greater than 75 years of age, unadjusted mortality was 5.1%, 13.5%, and 38.9%, respectively; ICU use was 29.8%, 35.2%, and 11.3%, respectively; and 30-day readmission was 11.6%, 8.8%, and 5.7%, respectively.

### Intensive Care Unit Admissions

Among patients admitted to the ICU (Table 5), those with COVID-19 were younger than those with influenza or other conditions (median age 59 years vs. 64 years and 66 years, respectively, SD=0.35), more likely to die (25.5% vs 19.9% and 11.9%, respectively, p<0.001) and had longer ICU LOS (median 10.9 days vs 6.0 days and 2.1 days, respectively p<0.001).

### Sensitivity Analyses

Results of all analyses were materially unchanged after excluding the 79 COVID-19 admissions without laboratory confirmation of virus infection (data not shown).

To avoid overestimating severity of illness because of interfacility transfers for critical care, we examined the subset of patients admitted from the ED. ED admissions (N=30,533) accounted for 70.3% of all admissions, including 79.8% of COVID-19 admissions, 86.0% of influenza admissions, and 69.7% of other admissions. Admissions for cardiac conditions and procedures were the most common causes of non-ED admissions. Findings were generally similar, except differences in hospital length-of-stay were somewhat attenuated and ICU use was less frequent among COVID-19 patients under 50 years (16.9%) and 50-75 years (24.9%) compared to the overall cohort (29.8% and 35.2%, respectively). Mortality among patients with COVID-19 under 50 years of age was also lower (<3.6% from ED and 5.0% overall). COVID-19 was consistently associated with worse outcomes than influenza and other conditions. See Appendix Tables 2-6 for details.

## Discussion

This multicentre cohort study describes patient characteristics, hospital resource use, clinical care, and outcomes for 1,027 adults hospitalized with COVID-19 in Ontario, Canada, during the first wave of the pandemic. To our knowledge, this represents the first detailed characterization of adult hospitalizations for COVID-19 in Canada. These findings can support future efforts to model hospital resource use as the pandemic evolves.^36^ Even after controlling for age, sex, comorbidity, and residence in long-term care, patients with COVID-19 had 3.5-times greater risk of death in hospital than patients with influenza or other conditions. Patients with COVID-19 were more likely to use ICU and invasive mechanical ventilation and had greater hospital length-of-stay.

Our study contributes to the emerging literature describing hospitalization for COVID-19. Two Canadian case series (involving 117 patients in Vancouver^6^ and 75 patients in Montreal^7^) described the care of ICU patients with COVID-19. In these reports, 57-63% of patients required invasive mechanical ventilation and 15-25% of patients died in hospital. Similarly, we found that invasive mechanical ventilation was used in 69% of ICU admissions for COVID-19 and 25% died. ICU care was received in 26% of all COVID-19 admissions, and 18% of those admitted through the ED. We extend this literature by including patients who did not require ICU and by reporting detailed data regarding presenting characteristics, clinical care, resource use, and outcomes. Mortality and ICU use in these Canadian hospitalizations were similar to the United States^1–3^, China^5,37^, and the United Kingdom^4^. We found that 8.7% of patients with COVID-19 were readmitted within 30 days, which is comparable to findings from the United States,^3,38^ and is important for modeling resource use.

Patients hospitalized with COVID-19 had a median age of 65 years, which was younger than in the United Kingdom (73 years)^4^ but somewhat older than studies in the United States and China (range 51-63 years)^1,2,5,37^. Patients with COVID-19 were more likely to be male (59%) and reside in long-term care (12%), which is consistent with evidence that COVID-19 affects men more severely^39^ and reflects the high burden of COVID-19 in long-term care facilities in Ontario^10,11^. However, 54% of patients hospitalized with COVID-19 had a Charlson comorbidity score of zero and 21% occurred in people under 50 years of age. Mortality in the younger group was relatively low (5.1% overall and <3.6% among ED admissions) but ICU use (29.8%) and 30-day readmission (11.6%) were high, reinforcing that COVID-19 can cause serious illness in younger people and those with relatively little comorbid disease.

Our study also contributes to comparisons of COVID-19 with seasonal influenza. The infection fatality rate of COVID-19 may be as much as 10-times greater than influenza^15,16^ but these comparisons are indirect and have been disputed.^13^ In a systematic review of observational studies, the in-hospital mortality rate of patients with influenza was reported to be approximately 3-6%^18^, which is similar to our finding (6.1%). After adjusting for age, sex, comorbidity and long-term care residence, compared to influenza, patients with COVID-19 had greater risk of death (3.5-times) and ICU admission (1.5-times), and longer hospital stay (1.4-times). Thus, COVID-19 is substantially more severe than seasonal influenza, and this information could assist with public health messaging.

More than 70% of patients hospitalized with COVID-19 received at least one physician order for respiratory-acting antibiotics, which was comparable to patients hospitalized with influenza. Bacterial co-infection is reported to affect up to 20-30% of ICU patients with influenza^40,41^ but only 8% of ICU patients with COVID-19.^42^ Our findings reinforce concerns that antibiotic use in COVID-19 may increase antimicrobial resistance and highlight the importance of future research in this area.

Our study has several limitations. First, we included 7 large academic hospitals. COVID-19 patients were transferred to these hospitals for critical care, which may lead us to overestimate the severity of COVID-19. To address this concern, we compared COVID-19 to influenza and other conditions, for which patients could also have been transferred. We also replicated our analyses in patients admitted through the ED to exclude inter-facility transfers. In this analysis, 20% of COVID-19 patients died in hospital, 18% required ICU, and the mortality rate compared to influenza was even greater (aRR 3.96). We believe our results are generalizable as mortality in our cohort was consistent with large studies from the United States^1^ and United Kingdom^4^ and we included approximately 25% of all COVID-19 hospitalizations in Ontario during the study period. Second, although COVID-19 disproportionately affects racial and ethnic minority communities^43^, data regarding patient race, ethnicity, language, or socioeconomic status is not collected systematically in Ontario hospitals and could not be reported. Third, we were unable to collect information about clinical history or symptoms, as this was not captured systematically in administrative or electronic medical record data. Fourth, we report 30-day readmission to a medical service or medical-surgical ICU at any participating hospital. Although this likely underestimates the total readmission rate, 82% of readmissions occur to the original hospital in our region^44^, and we were able to also capture readmissions to other participating hospitals. Fifth, the severity of seasonal influenza may vary from year-to-year and we compared COVID-19 only to influenza cases in 2019-2020. However, the mortality associated with influenza hospitalizations in our study (6.1%) is consistent with mortality rates of approximately 3-6% in a systematic review involving more than 120,000 influenza hospitalizations^18^, suggesting our results are likely generalizable. Finally, vital signs are not consistently recorded electronically at all participating hospitals, particularly when patients are in the emergency department or intensive care unit, which could have led to an underestimation of vital signs abnormalities.

## Conclusion

Adults hospitalized with COVID-19 at 7 hospitals in Ontario during the first wave of the pandemic used substantial hospital resources and suffered high rates of mortality. Patients hospitalized with COVID-19 had significantly greater mortality, ICU use, invasive mechanical ventilation use, and hospital length-of-stay than patients with influenza or other conditions.

## Data Availability

The data presented in this manuscript are available from the investigators upon reasonable request, and within the constraints placed by local REB approvals and data sharing agreements.

## Acknowledgments

We would like to acknowledge Dr. Radha Koppula for performing manual chart abstraction and Mr. Daniel Tamming for contributing to data analysis. We would also like to acknowledge the individuals and organizations that have made the data available for this research.

