## Supplementary material for "Patient characteristics, clinical care, resource use, and outcomes associated with hospitalization for COVID-19 in the Toronto area": Tables

Table 1. Characteristics of hospitalizations for COVID-19, influenza, and other conditions

| **Variable** | **COVID-19** | **Influenza** | **Other** | **SD^a^** |
| --- | --- | --- | --- | --- |
| Number of admissions | 1027 | 783 | 41652 |  |
| Number of unique patients | 972 (94.6) | 763 (97.4) | 33047 (79.3) | 0.47 |
| Age - median (IQR) | 65 (53, 79) | 68 (55, 80) | 68 (55, 80) | 0.07 |
| Age Group – N (%) |  |  |  | 0.09 |
| <50 | 218 (21.2) | 141 (18.0) | 8138 (19.5) |  |
| 50-75 | 480 (46.7) | 390 (49.8) | 19395 (46.6) |  |
| >75 | 329 (32.0) | 252 (32.2) | 14119 (33.9) |  |
| Female Gender – N (%) | 420 (40.9) | 385 (49.2) | 18292 (43.9) | 0.17 |
| Charlson Score – N (%) |  |  |  | 0.31 |
| 0 | 556 (54.1) | 304 (38.8) | 18168 (43.6) |  |
| 1 | 183 (17.8) | 175 (22.3) | 7409 (17.8) |  |
| 2+ | 288 (28.0) | 304 (38.8) | 16075 (38.6) |  |
| Hospital Frailty Risk Score – N (%) |  |  |  | 0.22 |
| Low (0-4) | 645 (62.8) | 562 (71.8) | 30272 (72.7) |  |
| Intermediate (5-14) | 344 (33.5) | 203 (25.9) | 10520 (25.3) |  |
| High (15+) | 38 (3.7) | 18 (2.3) | 860 (2.1) |  |
| Long-Term Care Resident – N (%) | 120 (11.7) | 35 (4.5) | 1202 (2.9) | 0.34 |
| Transfer from Acute Care Hospital – N (%) | 90 (8.8) | 24 (3.1) | 1937 (4.7) | 0.24 |
| Hypertension - N (%) | 356 (34.7) | 252 (32.2) | 14293 (34.3) | 0.05 |
| Diabetes Mellitus - N (%) | 284 (27.7) | 229 (29.2) | 10617 (25.5) | 0.05 |
| Renal failure - N (%) | 212 (20.6) | 169 (21.6) | 5856 (14.1) | 0.17 |
| Neurocognitive disorders - N (%) | 174 (16.9) | 105 (13.4) | 4205 (10.1) | 0.20 |
| Coronary heart disease - N (%) | 63 (6.1) | 63 (8.0) | 7595 (18.2) | 0.38 |
| Heart failure - N (%) | 62 (6.0) | 98 (12.5) | 4891 (11.7) | 0.22 |
| COPD - N (%) | 55 (5.4) | 96 (12.3) | 2366 (5.7) | 0.25 |

Table 1 Legend. Comparison of patients hospitalized with COVID-19, influenza, and all other conditions. a) SD = standardized difference. We calculated the standardized difference between COVID-19 and influenza and COVID-19 and other conditions, and report the largest of the two values. SD>0.1 reflects imbalance between groups. Comorbidities were categorized from ICD-10-CA discharge diagnoses using the CCSR tool. COPD: Chronic Obstructive Pulmonary Disease.

Table 2. Unadjusted clinical outcomes, resource use, and clinical care of patients with COVID-19, influenza, and other conditions

| **Variable** | **COVID-19** | **Influenza** | **Other** | **p** |
| --- | --- | --- | --- | --- |
| Number of Admissions | 1027 | 783 | 41652 |  |
| Death – N (%) | 204 (19.9) | 48 (6.1) | 2470 (5.9) | <0.001 |
| 7-day Readmission – N (%)^a^ | 37 (4.4) | 25 (3.3) | 1537 (3.9) | 0.553 |
| 30-day Readmission – N (%)^b^ | 66 (8.6) | 61 (8.2) | 3256 (8.8) | 0.805 |
| ICU – N (%) | 271 (26.4) | 141 (18.0) | 9769 (23.5) | <0.001 |
| Hospital LOS, days – median (IQR) | 8.7 (3.6, 18.9) | 4.8 (2.3, 10.4) | 4.4 (1.9, 8.9) | <0.001 |
| ED LOS, hours– median (IQR) | 8.7 (6.1, 13.2) | 21.1 (12.0, 32.2) | 13.6 (8.6, 24.1) | <0.001 |
| Invasive mechanical ventilation – N (%) | 190 (18.5) | 73 (9.3) | 4060 (9.7) | <0.001 |
| GI Endoscopy – N (%) | 21 (2.0) | 27 (3.4) | 3334 (8.0) | <0.001 |
| Bronchoscopy – N (%) | 21 (2.0) | 44 (5.6) | 711 (1.7) | <0.001 |
| Renal Replacement Therapy – N (%) | 79 (7.7) | 43 (5.5) | 1811 (4.3) | <0.001 |
| CT Thorax – N (%) | 202 (19.7) | 168 (21.5) | 6616 (15.9) | <0.001 |
| Respiratory Antibiotic – N (%) | 730 (71.6) | 599 (77.1) | 18981 (47.3) | <0.001 |
| Corticosteroid – N (%) | 170 (16.7) | 284 (36.6) | 8525 (21.3) | <0.001 |
| Warfarin or DOAC – N (%) | 157 (15.4) | 156 (20.1) | 8345 (20.8) | <0.001 |

Table 2 Legend. Readmission to medical service or medical-surgical ICU at any participating hospital is reported. For hospital resources and clinical care, we report the number of patients receiving at least one of the items described. a) After excluding patients who died and those discharged in the last 7 days of the study period, the denominator was 840 admissions for COVID-19, 748 for influenza, and 38939 for other. b) After excluding patients who died and those discharged in the last 30 days of the study period, the denominator was 768 admissions for COVID-19, 747 for influenza, and 36913 for other. ICU: Intensive Care Unit. LOS: Length-of-stay. GI: Gastrointestinal. Renal replacement therapy included hemodialysis and peritoneal dialysis and included both chronic use and new starts. CT: Computed tomography. Antibiotic: we report the use of respiratory-acting antibiotics (see Appendix). DOAC: direct-acting oral anticoagulant.

Table 3. Association between COVID-19 and clinical outcomes before and after multivariable adjustment

| **Outcome** | **Unadjusted** | | | | **Adjusted** | | | |
| --- | --- | --- | --- | --- | --- | --- | --- | --- |
|  | **COVID-19 vs Influenza** | | **COVID-19 vs Other** | | **COVID-19 vs Influenza** | | **COVID-19 vs Other** | |
|  | **Effect**  **(95% CI)** | **p** | **Effect**  **(95% CI)** | **p** | **Effect**  **(95% CI)** | **p** | **Effect**  **(95% CI)** | **p** |
| Death | 3.24 (2.40, 4.38) | <0.001 | 3.35 (2.94, 3.81) | <0.001 | 3.47 (2.57, 4.67) | <0.001 | 3.58 (3.16, 4.06) | <0.001 |
| ICU | 1.47 (1.22, 1.76) | <0.001 | 1.13 (1.01, 1.25) | 0.026 | 1.52 (1.27, 1.83) | <0.001 | 1.21 (1.09, 1.34) | <0.001 |
| Readmission | 1.05 (0.75, 1.47) | 0.764 | 0.97 (0.77, 1.23) | 0.826 | 1.01 (0.72, 1.42) | 0.964 | 0.94 (0.74, 1.19) | 0.609 |
| Hospital LOS | 1.31 (1.09, 1.58) | 0.003 | 1.66 (1.54, 1.79) | <0.001 | 1.40 (1.20, 1.64) | <0.001 | 1.79 (1.66, 1.92) | <0.001 |
| ICU LOS | 0.93 (0.57, 1.52) | 0.769 | 2.52 (2.20, 2.90) | <0.001 | 1.16 (0.83, 1.62) | 0.394 | 2.87 (2.44, 3.38) | <0.001 |

Table 3 Legend. Poisson regression models were fit for death, ICU, and readmission (effect: adjusted relative risk) and negative binomial regression models were fit for hospital LOS and ICU LOS (effect: adjusted rate ratio). Models were adjusted for patient age, sex, long-term care residence, Charlson comorbidity index score, and admitting hospital. Outcomes reported are: in-hospital death, admission to ICU at any point during hospitalization, readmission to a medical service or medical-surgical ICU at any participating hospital within 30-days of discharge, hospital LOS, and ICU LOS. ICU: intensive care unit. LOS: length-of-stay. Other: all admissions other than COVID-19 or influenza.

Table 4. Unadjusted outcomes stratified by age

| **Outcome** | **COVID-19** | **Influenza** | **Other** | **p** |
| --- | --- | --- | --- | --- |
| Age <50 years – N | 218 | 141 | 8138 |  |
| Death – N (%) | 11 (5.1) | <6 (<4.3)* | 178 (2.2) | 0.020 |
| 30-day Readmission – N (%) | 20 (11.6) | <6 (<4.3)* | 657 (9.1) | 0.046 |
| ICU – N (%) | 65 (29.8) | 27 (19.2) | 1795 (22.1) | 0.017 |
| Hospital LOS, days – median (IQR) | 5.6 (2.5, 12.9) | 3.5 (1.5, 8.2) | 3.3 (1.5, 6.9) | <0.001 |
| Age 50-75 years – N | 480 | 390 | 19395 |  |
| Death – N (%) | 65 (13.5) | 19 (4.9) | 931 (4.8) | <0.001 |
| 30-day Readmission – N (%) | 34 (8.8) | 27 (7.2) | 1491 (8.6) | 0.631 |
| ICU – N (%) | 169 (35.2) | 83 (21.3) | 5415 (27.9) | <0.001 |
| Hospital LOS, days – median (IQR) | 8.8 (3.6, 20.8) | 4.2 (2.0, 8.8) | 4.3 (1.8, 8.5) | <0.001 |
| Age >75 years – N | 329 | 252 | 14119 |  |
| Death – N (%) | 128 (38.9) | 26 (10.3) | 1361 (9.6) | <0.001 |
| 30-day Readmission – N (%) | 12 (5.7) | 29 (12.3) | 1108 (9.0) | 0.053 |
| ICU – N (%) | 37 (11.3) | 31 (12.3) | 2559 (18.1) | <0.001 |
| Hospital LOS, days – median (IQR) | 11.9 (4.8, 25.1) | 6.4 (3.7, 11.9) | 5.5 (2.5, 10.8) | <0.001 |

Table 4 Legend. 30-day readmission to medical service or medical-surgical ICU at any participating hospital is reported after excluding patients who died and those discharged in the last 30 days of the study period. ICU: Intensive Care Unit. LOS: Length-of-stay. *All counts less than 6 suppressed to reduce risk of patient reidentification.

Table 5. Characteristics, clinical outcomes, resource use, and clinical care of ICU patients

| **Variable** | **COVID-19** | **Influenza** | **Other** | **p** |
| --- | --- | --- | --- | --- |
| Number of admissions | 271 | 141 | 9769 |  |
| Age - median (IQR) | 59 (51, 70) | 64 (54, 74) | 66 (55, 76) | <0.001 |
| Age Group – N (%) |  |  |  | <0.001 |
| <50 | 65 (24.0) | 27 (19.1) | 1795 (18.4) |  |
| 50-75 | 169 (62.4) | 83 (58.9) | 5415 (55.4) |  |
| >75 | 37 (13.7) | 31 (22.0) | 2559 (26.2) |  |
| Female Gender – N (%) | 97 (35.8) | 61 (43.3) | 3535 (36.2) | 0.545 |
| Charlson Score – N (%) |  |  |  | <0.001 |
| 0 | 157 (57.9) | 46 (32.6) | 4619 (47.3) |  |
| 1 | 59 (21.8) | 30 (21.3) | 1874 (19.2) |  |
| 2+ | 55 (20.3) | 65 (46.1) | 3276 (33.5) |  |
| Hospital Frailty Risk Score – N (%) |  |  |  | <0.001 |
| Low (0-4) | 128 (47.2) | 66 (46.8) | 6888 (70.5) |  |
| Intermediate (5-14) | 135 (49.8) | 66 (46.8) | 2639 (27.0) |  |
| High (15+) | 8 (3.0) | 9 (6.4) | 242 (2.5) |  |
| Long-Term Care Resident – N (%) | 10 (3.7) | <6 (<4.3)* | 106 (1.1) | <0.001 |
| Transfer from Acute Care Hospital – N (%) | 84 (31.0) | 19 (13.5) | 1167 (11.9) | <0.001 |
| Death – N (%) | 69 (25.5) | 28 (19.9) | 1164 (11.9) | <0.001 |
| 7-day Readmission – N (%) | 8 (3.7) | 6 (4.8) | 286 (3.3) | 0.588 |
| 30-day Readmission – N (%) | 10 (5.0) | 14 (11.4) | 487 (5.8) | 0.03 |
| Hospital LOS, days – median (IQR) | 16.8 (10.6, 27.3) | 14.1 (6.8, 30.9) | 6.8 (3.5, 13.6) | <0.001 |
| ED LOS, hours – median (IQR) | 7.1 (4.6, 9.8) | 12.5 (7.2, 24.4) | 8.3 (4.3, 15.2) | <0.001 |
| ICU LOS, days – median (IQR) | 10.9 (4.0, 17.8) | 6.0 (2.3, 13.0) | 2.1 (1.1, 4.6) | <0.001 |
| Invasive mechanical ventilation – N (%) | 188 (69.4) | 73 (51.8) | 3923 (40.2) | <0.001 |
| GI Endoscopy – N (%) | 7 (2.6) | 13 (9.2) | 596 (6.1) | 0.016 |
| Bronchoscopy – N (%) | 19 (7.0) | 33 (23.4) | 381 (3.9) | <0.001 |
| Renal Replacement Therapy – N (%) | 52 (19.2) | 18 (12.8) | 770 (7.9) | <0.001 |
| CT Thorax – N (%) | 77 (28.4) | 53 (37.6) | 1828 (18.7) | <0.001 |
| Antibiotic – N (%) | 244 (90.7) | 128 (92.1) | 5600 (57.9) | <0.001 |
| Corticosteroid – N (%) | 89 (33.1) | 69 (49.6) | 2110 (21.8) | <0.001 |
| Warfarin or DOAC – N (%) | 47 (17.5) | 34 (24.5) | 2153 (22.3) | 0.141 |

Table 5 Legend. Readmission to medical service or medical-surgical ICU at any participating hospital is reported. For hospital resources and clinical care, we report the number of patients receiving at least one of the item described. ICU: Intensive Care Unit. LOS: Length-of-stay. GI: Gastrointestinal. Renal replacement therapy included hemodialysis and peritoneal dialysis and included both chronic use and new starts. CT: Computed tomography. Antibiotic: we report the use of respiratory-acting antibiotics (see Appendix). DOAC: direct-acting oral anticoagulant. *Exact number suppressed to reduce risk of patient reidentification.

**Appendix.** Respiratory-acting antibiotics

We report the use of antibiotics with action against respiratory bacterial pathogens, as described in clinical practice guidelines and prior literature.^23-25^

**Antibiotic classes included**

Third Generation Cephalosporin

Macrolide

Fluoroquinolone

Penicillin-derived beta-lactams/beta-lactamase

Other

Action against methicillin-resistant S. aureus

Simple Penicillins

Ceftazidime

Tetracyclines

Carbapenem (with and without pseudomonas coverage)

Clindamycin

**Specific antibiotics included**
Third generation cephalosporin=Ceftriaxone, cefotaxime, cefepime, cefdinir, cefditoren, cefpoxidime, ceftaroline. Macrolide=Azithromycin, clarithromycin, erythromycin, streptomycin. Fluoroquinolone=levofloxacin, moxifloxacin, ciprofloxacin, gemifloxicin. Tetracyclines=doxycycline. Penicillin-derived beta-lactam/beta-lactamases=amoxicillin-clavulinic acid, ampicillin-sulbactam, ticarcillin-clavulanate, piperacillin-tazobactam. Carbapenems=meropenem, imipenem, impenem+cilastatin. ertapenem. Action against methicillin-resistant S. aureus =vancomycin, linezolid. Simple Penicillins=Penicillin G, amoxicillin, ticarcillin, flucloxacillin, ampicillin, piperacillin.  Other=aztreonam, colistin, gentamicin, trimethoprim-sulfamethoxazole, first and second generation cephalosporins (cefazolin, cefprozil, cefuroxime, cephalexin).

Appendix Table 1. Presenting vital signs and laboratory test results in patients hospitalized with COVID-19, Influenza, and Other Conditions.

| **Variable** | **Result** | **Number Performed** | **Result** | **Number Performed** | **Result** | **Number Performed** | **p*** |
| --- | --- | --- | --- | --- | --- | --- | --- |
| Temperature - median (IQR) | 36.9 (36.6, 37.5) | 863 (84.0) | 36.9 (36.6, 37.4) | 557 (71.1) | 36.7 (36.4, 36.9) | 27720 (66.4) | <0.001 |
| Systolic BP - median (IQR) | 119 (110, 136) | 870 (84.7) | 110 (110, 136) | 540 (69.0) | 113 (110, 136) | 26844 (64.3) | 0.139 |
| Diastolic BP - median (IQR) | 69 (66, 79) | 869 (84.6) | 66 (66, 76) | 540 (69.0) | 66 (66, 77) | 26845 (64.3) | 0.004 |
| Heart Rate - median (IQR) | 87 (75, 100) | 869 (84.6) | 88 (76, 99) | 540 (69.0) | 82 (70, 95) | 26858 (64.3) | <0.001 |
| Resp Rate - median (IQR) | 19 (18, 20) | 805 (78.4) | 18 (18, 20) | 488 (62.3) | 18 (18, 20) | 24081 (57.7) | <0.001 |
| S/F ratio - median (IQR) | 452 (424, 467) | 666 (64.8) | 452 (431, 467) | 427 (54.5) | 462 (452, 471) | 22003 (52.7) | <0.001 |
| Supplemental O_2_ - N (%) | 367 (46.0) | 798 (77.7) | 189 (40.1) | 471 (60.2) | 5107 (21.3) | 24058 (57.6) | <0.001 |
| Hemoglobin - median (IQR) | 124 (108, 138) | 936 (91.1) | 120 (105, 135) | 753 (96.2) | 120 (102, 136) | 36396 (87.4) | <0.001 |
| WBC Count -median (IQR) | 7.5 (5.4, 10.6) | 935 (91.0) | 7.5 (5.2, 10.5) | 753 (96.2) | 8.6 (6.4, 11.9) | 36382 (87.4) | 0.001 |
| Plt Count - median (IQR) | 212 (163, 277) | 932 (90.7) | 180 (141, 235) | 751 (95.9) | 216 (163, 280) | 36347 (87.3) | 0.097 |
| Sodium - median (IQR) | 137 (133, 140) | 970 (94.4) | 136 (133, 139) | 773 (98.7) | 138 (135, 140) | 37376 (89.8) | <0.001 |
| Creatinine - median (IQR) | 87 (68, 122) | 932 (90.7) | 92 (69, 131) | 748 (95.5) | 89 (69, 126) | 36340 (87.3) | 0.371 |
| Lactate - median (IQR) | 1.8 (1.3, 2.4) | 720 (70.1) | 1.8 (1.3, 2.5) | 602 (76.9) | 1.8 (1.3, 2.7) | 19089 (46.0) | <0.001 |
| D-dimer - median (IQR) | 1020 (682, 1898) | 394 (38.4) | 993 (441, 1821) | 34 (4.3) | 1030 (519, 1986) | 1539 (3.7) | 0.215 |
| Arterial pCO2 - median (IQR) | 44 (36, 54) | 208 (20.3) | 42 (34, 49) | 112 (14.3) | 40 (35, 46) | 3998 (9.6) | <0.001 |
| Arterial pO2 - median (IQR) | 74 (62, 94) | 208 (20.3) | 82 (66, 128) | 112 (14.3) | 108 (80, 158) | 4000 (9.6) | <0.001 |
| Venous pCO2 - median (IQR) | 43 (38, 49) | 462 (45.0) | 44 (39, 51) | 402 (51.3) | 44 (38, 51) | 10169 (24.3) | <0.001 |

Appendix Table 1 Legend. Laboratory test results and vital signs collected between emergency department triage and 48 hours after admission. We report the first result for each patient. Vital signs are not consistently recorded electronically at all hospitals, particularly when patients are in the emergency department or an intensive care unit. Vital signs were not electronically extracted from one participating hospital. For that hospital, vital signs were manually abstracted only for patients with COVID-19. *p-value is reported for the difference in vital/test results across the 3 groups. Temperature is reported in degrees census. BP = Blood Pressure, and is reported in mmHg. Heart rate and resp (respiratory) rate are reported in counts per minute. S/F ratio is the ration of oxygen saturation to fraction of inspired oxygen. Supplemental O_2_ is the number of patients who required any amount of supplemental oxygen or mechanical ventilation in the observation window (between ED triage and 48 hours after admission). Hemoglobin: g/L. WBC: White Blood Cell count x10^9^/L. Plt: Platelet Count x10^9^/L. Sodium: mmol/L. Creatinine: umol/L. Lactate: mmol/L. D-dimer ug/L (values standardized to this unit across sites if measurement units differed). pCO2 and pO2: mmHg.

Appendix Table 2. Characteristics of hospitalizations for COVID-19, influenza, and other conditions, among patients admitted from the emergency department

| **Variable** | **COVID-19** | **Influenza** | **Other** | **SD^a^** |
| --- | --- | --- | --- | --- |
| Number of admissions | 753 | 673 | 29107 |  |
| Number of unique patients | 727 (96.5) | 660 (98.1) | 23513 (80.8) |  |
| Age - median (IQR) | 66 (53, 80) | 69 (56, 81) | 69 (55, 82) | 0.09 |
| Age Group – N (%) |  |  |  | 0.14 |
| <50 | 166 (22.0) | 115 (17.1) | 5829 (20.0) |  |
| 50-75 | 333 (44.2) | 335 (49.8) | 12530 (43.0) |  |
| >75 | 254 (33.7) | 223 (33.1) | 10748 (36.9) |  |
| Female Gender – N (%) | 316 (42.0) | 334 (49.6) | 13442 (46.2) | 0.15 |
| Charlson Score – N (%) |  |  |  | 0.31 |
| 0 | 406 (53.9) | 259 (38.5) | 12004 (41.2) |  |
| 1 | 126 (16.7) | 146 (21.7) | 4855 (16.7) |  |
| 2+ | 221 (29.3) | 268 (39.8) | 12248 (42.1) |  |
| Hospital Frailty Risk Score – N (%) |  |  |  | 0.13 |
| Low (0-4) | 500 (66.4) | 484 (71.9) | 19759 (67.9) |  |
| Intermediate (5-14) | 227 (30.1) | 174 (25.9) | 8638 (29.7) |  |
| High (15+) | 26 (3.5) | 15 (2.2) | 710 (2.4) |  |
| Long-Term Care Resident – N (%) | 108 (14.3) | 33 (4.9) | 1091 (3.7) | 0.38 |
| Transfer from Acute Care Hospital – N (%) | 6 (0.8) | <6 (<0.9)* | 208 (0.7) | 0.07 |
| Hypertension - N (%) | 257 (34.1) | 222 (33.0) | 9872 (33.9) | 0.02 |
| Diabetes Mellitus - N (%) | 206 (27.4) | 194 (28.8) | 7413 (25.5) | 0.04 |
| Renal failure - N (%) | 139 (18.5) | 136 (20.2) | 4625 (15.9) | 0.07 |
| Neurocognitive disorders - N (%) | 122 (16.2) | 92 (13.7) | 3361 (11.5) | 0.13 |
| Coronary heart disease - N (%) | 55 (7.3) | 52 (7.7) | 3486 (12.0) | 0.16 |
| Heart failure - N (%) | 48 (6.4) | 82 (12.2) | 3677 (12.6) | 0.21 |
| COPD - N (%) | 47 (6.2) | 80 (11.9) | 1986 (6.8) | 0.20 |

Appendix Table 2 Legend. Comparison of patients hospitalized with COVID-19, influenza, and all other conditions. a) SD = standardized difference. We calculated the standardized difference between COVID-19 and influenza and COVID-19 and other conditions, and report the largest of the two values. SD>0.1 reflects imbalance between groups. Comorbidities were categorized from ICD-10-CA discharge diagnoses using the CCSR tool. COPD: Chronic Obstructive Pulmonary Disease. *All counts less than 6 suppressed to reduce risk of patient reidentification.

Appendix Table 3. Unadjusted clinical outcomes, resource use, and clinical care of patients with COVID-19, influenza, and other conditions, among patients admitted from the emergency department

| **Variable** | **COVID-19** | **Influenza** | **Other** | **p** |
| --- | --- | --- | --- | --- |
| Number of Admissions | 753 | 673 | 29107 |  |
| Death – N (%) | 154 (20.5) | 37 (5.5) | 1872 (6.4) | <0.001 |
| 7-day Readmission – N (%)^a^ | 27 (4.4) | 21 (3.2) | 1142 (4.2) | 0.459 |
| 30-day Readmission – N (%)^b^ | 49 (8.7) | 53 (8.2) | 2621 (10.2) | 0.14 |
| ICU – N (%) | 137 (18.2) | 101 (15.0) | 4906 (16.9) | 0.272 |
| Hospital LOS, days – median (IQR) | 6.8 (3.0, 16.2) | 4.7 (2.2, 9.5) | 4.6 (2.3, 9.3) | <0.001 |
| ED LOS, hours– median (IQR) | 8.7 (6.1, 13.2) | 21.1 (12.0, 32.2) | 13.6 (8.6, 24.1) | <0.001 |
| Invasive mechanical ventilation – N (%) | 79 (10.5) | 50 (7.4) | 1636 (5.6) | <0.001 |
| GI Endoscopy – N (%) | 16 (2.1) | 23 (3.4) | 2678 (9.2) | <0.001 |
| Bronchoscopy – N (%) | <6 (<0.8)* | 32 (4.8) | 487 (1.7) | <0.001 |
| Renal Replacement Therapy – N (%) | 29 (3.9) | 37 (5.5) | 1079 (3.7) | 0.053 |
| CT Thorax – N (%) | 151 (20.1) | 147 (21.8) | 5277 (18.1) | 0.021 |
| Respiratory Antibiotic – N (%) | 518 (69.4) | 505 (75.5) | 13912 (48.1) | <0.001 |
| Corticosteroid – N (%) | 104 (13.9) | 243 (36.3) | 6302 (21.8) | <0.001 |
| Warfarin or DOAC – N (%) | 111 (14.9) | 133 (19.9) | 6030 (20.8) | <0.001 |

Appendix Table 3 Legend. Readmission to medical service or medical-surgical ICU at any participating hospital is reported. For hospital resources and clinical care, we report the number of patients receiving at least one of the items described. ICU: Intensive Care Unit. a) After excluding patients who died and those discharged in the last 7 days of the study period, the denominator was 610 admissions for COVID-19, 647 for influenza, and 27092 for other. b) After excluding patients who died and those discharged in the last 30 days of the study period, the denominator was 562 admissions for COVID-19, 646 for influenza, and 25741 for other. LOS: Length-of-stay. GI: Gastrointestinal. Renal replacement therapy included hemodialysis and peritoneal dialysis and included both chronic use and new starts. CT: Computed tomography. Antibiotic: we report the use of respiratory-acting antibiotics (see Appendix). DOAC: direct-acting oral anticoagulant. *All counts less than 6 suppressed to reduce risk of patient reidentification.

Appendix Table 4. Association between COVID-19 and clinical outcomes before and after multivariable adjustment, among patients admitted from the emergency department

| **Outcome** | **Unadjusted** | | | | **Adjusted** | | | |
| --- | --- | --- | --- | --- | --- | --- | --- | --- |
|  | **COVID-19 vs Influenza** | | **COVID-19 vs Other** | | **COVID-19 vs Influenza** | | **COVID-19 vs Other** | |
|  | **Effect**  **(95% CI)** | **p** | **Effect**  **(95% CI)** | **p** | **Effect**  **(95% CI)** | **p** | **Effect**  **(95% CI)** | **p** |
| Death | 3.72 (2.64, 5.25) | <0.001 | 3.18 (2.74, 3.69) | <0.001 | 3.96 (2.82, 5.57) | <0.001 | 3.43 (2.98, 3.95) | <0.001 |
| ICU | 1.21 (0.96, 1.53) | 0.109 | 1.08 (0.93, 1.26) | 0.33 | 1.28 (1.00, 1.63) | 0.047 | 1.13 (0.97, 1.32) | 0.128 |
| Readmission | 1.06 (0.73, 1.54) | 0.749 | 0.86 (0.65, 1.12) | 0.261 | 1.08 (0.73, 1.58) | 0.702 | 0.84 (0.64, 1.10) | 0.21 |
| Hospital LOS | 1.19 (0.96, 1.47) | 0.12 | 1.42 (1.29, 1.56) | <0.001 | 1.18 (0.99, 1.41) | 0.071 | 1.51 (1.38, 1.66) | <0.001 |
| ICU LOS | 0.64 (0.35, 1.17) | 0.15 | 1.69 (1.38, 2.06) | <0.001 | 0.75 (0.48, 1.17) | 0.208 | 1.73 (1.38, 2.16) | <0.001 |

Appendix Table 4 Legend. Poisson regression models were fit for death, ICU, and readmission (effect: adjusted relative risk) and negative binomial regression models were fit for hospital LOS and ICU LOS (effect: adjusted rate ratio). Models were adjusted for patient age, sex, long-term care residence, Charlson comorbidity index score, and admitting hospital. Outcomes reported are: in-hospital death, admission to ICU at any point during hospitalization, readmission to a medical service or medical-surgical ICU at any participating hospital within 30-days of discharge, hospital LOS, and ICU LOS. ICU: intensive care unit. LOS: length-of-stay. Other: all admissions other than COVID-19 or influenza.

Appendix Table 5. Unadjusted outcomes stratified by age, among patients admitted from the emergency department

| **Outcome** | **COVID-19** | **Influenza** | **Other** | **p** |
| --- | --- | --- | --- | --- |
| Age <50 years – N | 166 | 115 | 5829 |  |
| Death – N (%) | <6 (<3.6)* | <6 (<5.2)* | 125 (2.1) | 0.623 |
| 30-day Readmission – N (%) | 14 (10.3) | <6 (<5.2)* | 567 (10.9) | 0.048 |
| ICU – N (%) | 28 (16.9) | 15 (13.0) | 1048 (18.0) | 0.370 |
| Hospital LOS, days – median (IQR) | 4.7 (1.8, 9.0) | 3.2 (1.3, 6.7) | 3.2 (1.6, 6.6) | 0.003 |
| Age 50-75 years – N | 333 | 335 | 12530 |  |
| Death – N (%) | 42 (12.6) | 13 (3.9) | 654 (5.2) | <0.001 |
| 30-day Readmission – N (%) | 27 (9.9) | 23 (7.1) | 1136 (10.2) | 0.192 |
| ICU – N (%) | 83 (24.9) | 60 (17.9) | 2392 (19.1) | 0.024 |
| Hospital LOS, days – median (IQR) | 6.7 (2.9, 16.4) | 4.4 (1.9, 8.4) | 4.4 (2.3, 8.8) | <0.001 |
| Age >75 years – N | 254 | 223 | 10748 |  |
| Death – N (%) | 108 (42.5) | 23 (10.3) | 1093 (10.2) | <0.001 |
| 30-day Readmission – N (%) | 8 (5.2) | 26 (12.4) | 918 (9.8) | 0.071 |
| ICU – N (%) | 26 (10.2) | 26 (11.7) | 1466 (13.6) | 0.209 |
| Hospital LOS, days – median (IQR) | 10.6 (4.5, 24.0) | 6.0 (3.7, 11.7) | 5.7 (2.9, 11.0) | <0.001 |

Appendix Table 5 Legend. Readmission to medical service or medical-surgical ICU at any participating hospital is reported. ICU: Intensive Care Unit. LOS: Length-of-stay. *All counts less than 6 suppressed to reduce risk of patient reidentification.

Appendix Table 6. Characteristics, clinical outcomes, resource use, and clinical care of ICU patients, among patients admitted from the emergency department

| **Variable** | **COVID-19** | **Influenza** | **Other** | **p** |
| --- | --- | --- | --- | --- |
| Number of admissions | 137 | 101 | 4906 |  |
| Age - median (IQR) | 62 (52, 73) | 67 (56, 77) | 66 (53, 78) | 0.050 |
| Age Group – N (%) |  |  |  | 0.008 |
| <50 | 28 (20.4) | 15 (14.9) | 1048 (21.4) |  |
| 50-75 | 83 (60.6) | 60 (59.4) | 2392 (48.8) |  |
| >75 | 26 (19.0) | 26 (25.7) | 1466 (29.9) |  |
| Male Gender – N (%) | 78 (56.9) | 56 (55.4) | 2990 (60.9) | 0.349 |
| Charlson Score – N (%) |  |  |  | <0.001 |
| 0 | 73 (53.3) | 30 (29.7) | 2163 (44.1) |  |
| 1 | 31 (22.6) | 23 (22.8) | 771 (15.7) |  |
| 2+ | 33 (24.1) | 48 (47.5) | 1972 (40.2) |  |
| Hospital Frailty Risk Score – N (%) |  |  |  | 0.053 |
| Low (0-4) | 78 (56.9) | 44 (43.6) | 2849 (58.1) |  |
| Intermediate (5-14) | 54 (39.4) | 50 (49.5) | 1864 (38.0) |  |
| High (15+) | <6 (<4.4)* | 7 (6.9) | 193 (3.9) |  |
| Long-Term Care Resident – N (%) | 8 (5.8) | <6 (<6.0)* | 100 (2.0) | 0.002 |
| Death – N (%) | 41 (29.9) | 19 (18.8) | 812 (16.6) | <0.001 |
| 30-day Readmission – N (%) | 6 (5.9) | 10 (11.1) | 299 (7.4) | 0.344 |
| Hospital LOS, days – median (IQR) | 15.9 (9.7, 26.5) | 15.0 (7.3, 32.6) | 8.2 (3.9, 16.7) | <0.001 |
| ED LOS, hours – median (IQR) | 7.1 (4.6, 9.8) | 12.5 (7.2, 24.4) | 8.3 (4.3, 15.2) | <0.001 |
| ICU LOS, days – median (IQR) | 7.9 (2.7, 15.4) | 6.3 (2.8, 13.3) | 2.8 (1.5, 5.8) | <0.001 |
| Invasive mechanical ventilation – N (%) | 77 (56.2) | 50 (49.5) | 1536 (31.3) | <0.001 |
| GI Endoscopy – N (%) | <6 (<4.4)* | 11 (10.9) | 386 (7.9) | 0.098 |
| Bronchoscopy – N (%) | <6 (<4.4)* | 22 (21.8) | 229 (4.7) | <0.001 |
| Renal Replacement Therapy – N (%) | 19 (13.9) | 13 (12.9) | 378 (7.7) | <0.001 |
| CT Thorax – N (%) | 43 (31.4) | 41 (40.6) | 1228 (25.0) | 0.001 |
| Antibiotic – N (%) | 119 (88.1) | 94 (94.0) | 2881 (59.0) | <0.001 |
| Corticosteroid – N (%) | 40 (29.6) | 48 (48.0) | 1282 (26.3) | <0.001 |
| Warfarin or DOAC – N (%) | 24 (17.8) | 25 (25.0) | 1117 (22.9) | 0.328 |

Appendix Table 6 Legend. Readmission to medical service or medical-surgical ICU at any participating hospital is reported. For hospital resources and clinical care, we report the number of patients receiving at least one of the items described. ICU: Intensive Care Unit. LOS: Length-of-stay. GI: Gastrointestinal. Renal replacement therapy included hemodialysis and peritoneal dialysis and included both chronic use and new starts. CT: Computed tomography. Antibiotic: we report the use of respiratory-acting antibiotics (see Appendix). DOAC: direct-acting oral anticoagulant. *Exact number suppressed to reduce risk of patient reidentification.
